# A Diffusion tensor imaging study to compare normative fractional anisotropy values with patients suffering from Parkinson’s disease in the brain grey and white matter

**DOI:** 10.1101/2020.06.09.20126755

**Authors:** Rahul P Kotian, K Prakashini, N Sreekumaran Nair

**Author notes:** Corresponding author: Dr. Rahul P Kotian, Professor, (PhD), MSc. Medical Imaging Technology, Department of Medical Imaging Technology, College of Allied Health Sciences, Srinivas University, Mukka Karnataka, India.,. SCOPUS id: 56073838900.

## Abstract

**Background:** Diffusion tensor imaging (DTI) appears as a sensitive method to study Parkinson’s disease (PD) pathophysiology and severity. Fractional anisotropy (FA) value is one of the scalar derivatives of DTI used to find out anisotropy within a voxel in a tissue and used for determining white matter integrity in aging and neurodegenerative diseases. We studied DTI derived FA in early PD subjects as their routine MRI scans were normal.

**Methods:** 40 patients with early PD and 40 healthy controls were employed to evaluate changes in microstructural white and grey matter in the brain’s using DTI derived FA values. Comparison of FA values in the brain’s white and grey matter of patients with PD and age matched controls at the corpus callosum, centrum semiovale, pons, putamen, caudate nucleus, substantia nigra, cerebral peduncles and cerebellar peduncles, was done using a region of interest (ROI) technique, with b-value 1000s/mm^2^ and TE=100 milliseconds using 1.5T MRI system.

**Results:** PD patients showed differences in FA values in both the grey and white matter areas of the brain’s compared to healthy controls. Our study revealed the presence of damage in the substantia nigra, corpus callosum, putamen and cerebral peduncles mainly in the PD group.

**Conclusion:** Our findings indicate that DTI and region of interest (ROI) methods can be used in patients with early PD to study microstructural alterations mainly in the substantia nigra, putamen and corpus callosum.

## 1. Introduction

Parkinson’s disease (PD) is a dopamine related dysfunction and its diagnosis is made in the presence of at least one motor systems, such as, bradykinesia, rest tremor, rigidity and postural instability [1]. PD manifests within the brain very slowly and hence conventional MRI scans fail to pick any significant changes. However, MRI brain scans can show age related and atrophy changes but the overall diagnostic value in PD is poor. Early diagnosis in PD allows the clinicians to improve the quality of life and reduce morbidity of these PD patients.

The search of a neuroimaging biomarker for early PD diagnosis is crucial and might have a high impact on a patient’s quality of life. Diffusion tensor imaging (DTI) is a beneficial tool to assess white and grey matter abnormalities, which can be used in the long-term assessment of the patients and serve as the imaging markers of disease progression and treatment response [2, 3]. The measures obtained from DTI are fractional anisotropy (FA), mean diffusivity (MD), radial diffusivity (RD) and axial diffusivity (AD). FA quantifies the preferred diffusion direction of water molecules through white matter tracts, and MD represents diffusion magnitude [4, 5]. FA is one of the most commonly used quantitative measure of diffusion in the brain in PD cases. The recent trend in using DTI as a diagnostic tool in PD has generated importance on our understanding of structural abnormalities of this disease and the robust technique for PD imaging has been the use of FA [6].

Previous studies in DTI still lack vital information about alteration beyond the substantia nigra (SN), according to a meta-analysis related degeneration in PD [7]. Recently, DTI has been proved to be useful in PD detection in both grey and white matter regions of the brain [8,9]. Some studies also focused on subcortical, cortical, white matter, cerebellar regions, and reported significant alterations in PD in many regions including the substantia nigra, caudate nucleus, putamen, globus pallidus, olfactory cortex and white matter of the corpus callosum [10]. Putaminal diffusivity correlates with disease progression in Parkinson’s Disease which shows involvement in brain grey matter [11].

Changes in FA values has been reported in numerous cases on PD, but its clinical use in diagnosis and treatment remains challenging and has not been adopted till now [12-16]. Tract based spatial statistics (TBSS) studies published till date reported a higher rate of false negatives and do not suggest a definite diagnosis and correlation [17]. Inclusion of small set of ROIs’ in studies reported on PD in both grey and white matter have led to unclear findings about FA on PD [18].

The main purpose of this study was to investigate the role of DTI imaging in different grey and white matter regions of the brain using FA values with ROI technique involved in early PD among those of healthy controls. We considered freehand ROI, as a unique technique for calculating FA at the grey and white regions of the brain because we were able to select the exact size, location and position of the desired anatomy to be evaluated unlike TBSS techniques.

## 2. Methods

The study protocol followed was reviewed and approved by the Research Committee of Manipal College of Health Profession and Manipal Academy of Higher Education, and ethical clearance was also obtained by Kasturba Medical College and Hospital, Manipal Academy of Higher Education, Manipal. A detailed explanation about the study was given by the principal investigator after which they provided consent for publication. All the patients included in this research gave written informed consent to publish the data contained within this study.

### 2.1 Subjects

This was a case-control study with a prospective design. A total of 40 patients with PD aged 53–75 years (25 males, 15 females, mean age: 64.05±8.79 years); and healthy control subjects (25 males, 15 females, mean age: 65.05±8.37 years) were enrolled. The duration of the disease was < 1 years in 25 cases and 1–3 years in 15 cases. All patients met the UK PD Society Brain Bank Clinical Diagnostic Criteria for probable PD. The Unified Parkinson Disease Rating scale (UPDRS) was also taken into consideration.

All the controls underwent a rigorous screening for neurological abnormalities including a routine MR brain imaging. Patients with clinically diagnosed parkinsonian syndromes i.e. early PD (0-3 years), were included in the study in the age group of 40-75 years. Patients suffering from vascular parkinsonism, multiple system atrophy (MSA) with (PD), PD dementia (PDD), or Lewy body dementia were excluded from the study. They were also excluded in the presence of the following criteria: 1) presence of structural brain abnormalities, 2) history of intracranial surgery or 3) major physical or neuropsychiatric disorders. Subjects with mild atrophy were considered eligible for the study.

### 2.2 Image Acquisition

Images were acquired on a 1.5-Tesla Philips Achieva, class IIA series using a 16 channel head coil system. Conventional imaging (T2-weighted FLAIR axial and T1-weighted sagittal sequences) was performed to rule out neurological abnormalities. DTI images were acquired using a single shot echo-planar imaging (EPI) sequence with the following parameters: b-value = 1000, TR/TE/slice thickness = 8648ms/100ms/2 mm, matrix size = 112 × 110, and FOV =224 × 224 mm. Parallel imaging was used, with an acceleration factor of two and generalized auto calibrating partially parallel acquisition (GRAPPA) reconstruction and EPI factor of 55. Diffusion-weighting gradients were applied in 15 non-collinear directions.

### 2.3 DTI derived measures from ROIs

A ROI technique was used to estimate FA values. The following ROIs were included in our analyses: corpus callosum, centrum semiovale, pons, substantia nigra, thalamus, cerebral peduncles, cerebellar peduncles, caudate nucleus and putamen. We used a voxel size of eight for corpus callosum, centrum semiovale and pons, a 4 voxel ROI at the cerebellar peduncles; while a single voxel was used to measure ROI’s in the rest of the regions as depicted in Fig. 1 – 6.

**Fig. 1.**
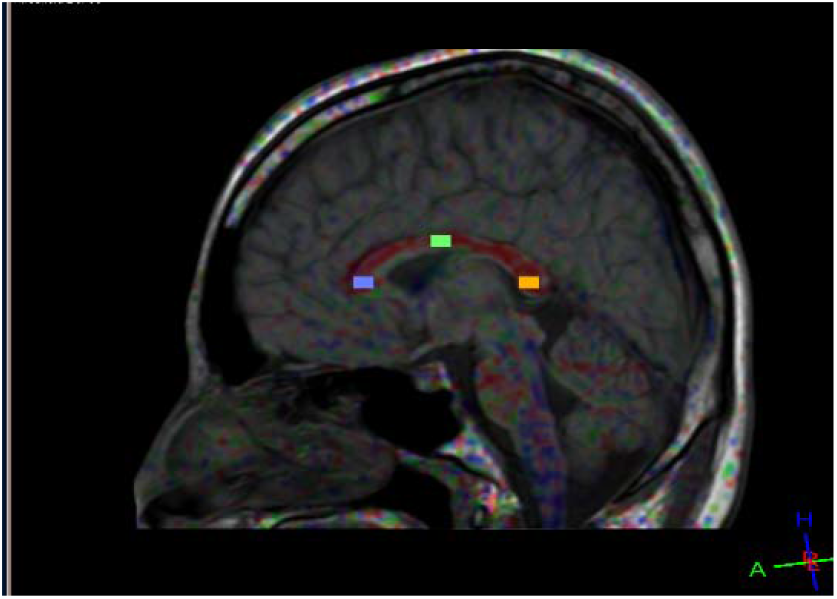
ROI at Corpus callosum- (genu, body and splenium)

**Fig. 2.**
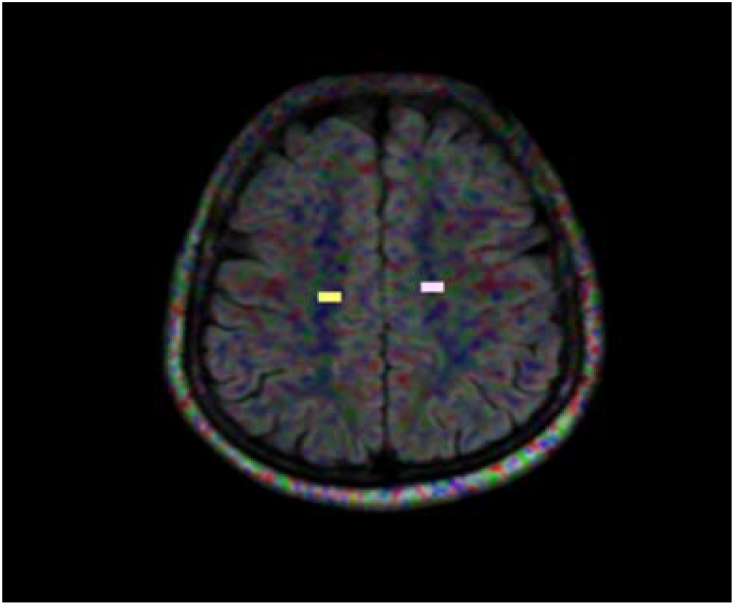
ROI at centrum semiovale (Right and Left)

**Fig. 3.**
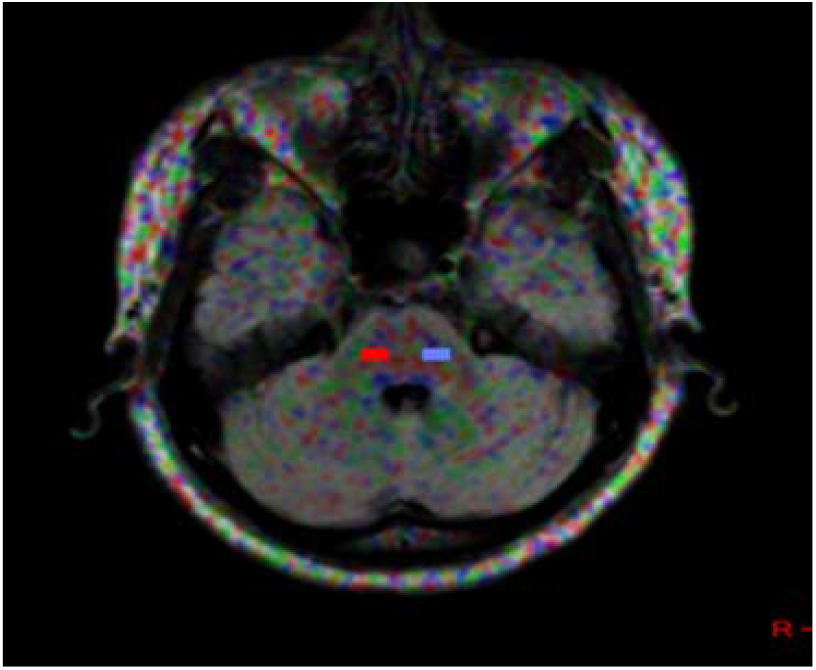
ROI at Pons- (Right and Left)

**Fig. 4.**
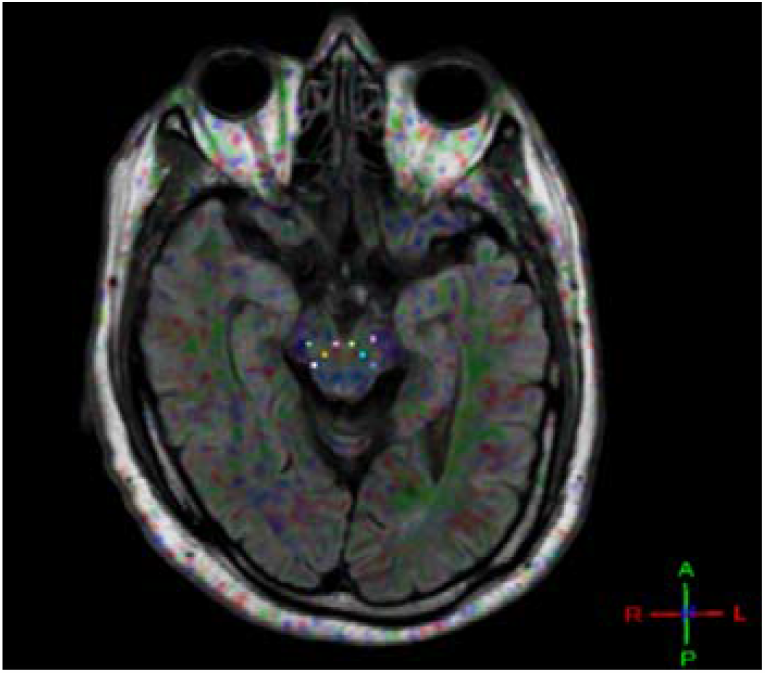
ROI at Substantia nigra (Right & Left) And Cerebral Peduncles (Right & Left)

**Fig. 5.**
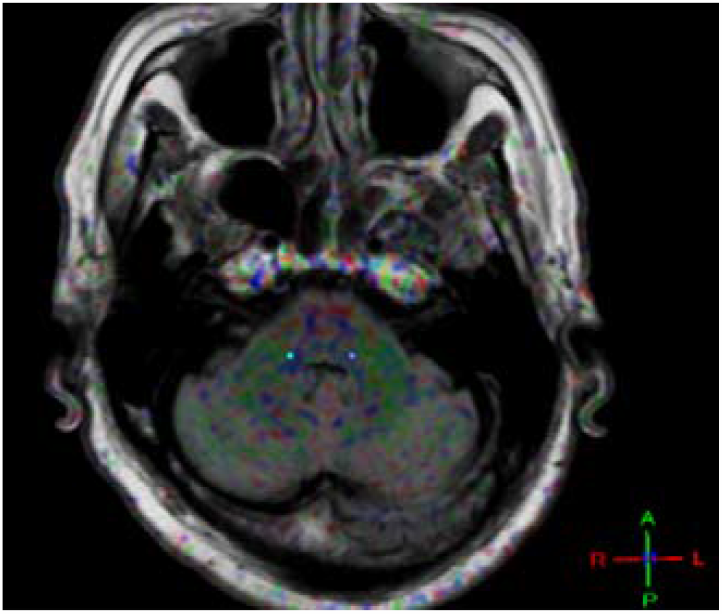
ROI at Cerebellar Peduncles (Right & Left)

**Fig. 6.**
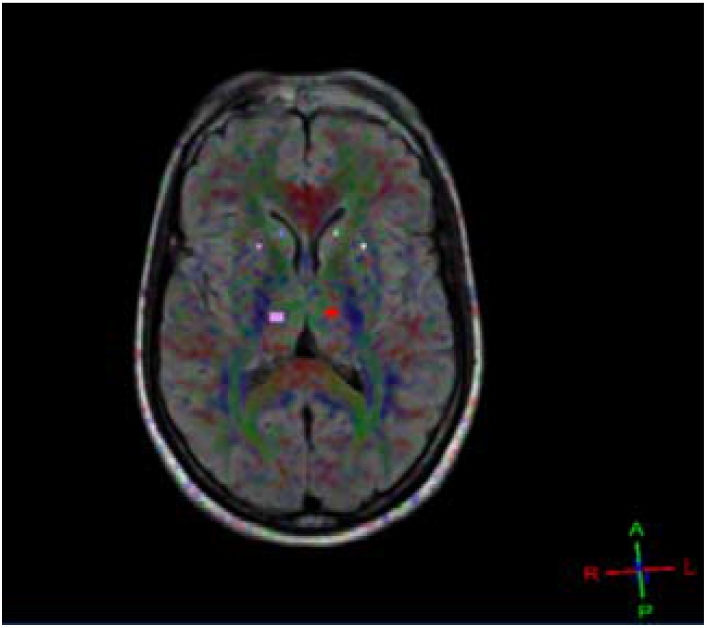
ROI at Caudate Nucleus (Right & Left), Putamen (Right & Left), Thalamus (Right & Left)

### 2.4 Image Analysis & Post-processing

ROI identification and estimation of FA values were done using the Philips extended workspace. The DTI image sets were loaded along with sequence Flair axial and T1 Sagittal for anatomy correlation to visualize and calculate FA values. ROI’s using standard techniques and anatomy reference overlap sequences were drawn in the brain regions mentioned above using DTI Philips extended MR fiber track software version 7.1.5.1.

### 2.5 Statistical Analysis

All analyses were performed using the SPSS software package (version 16.0). The independent sample t-test was used to compare the difference in FA values between PD patients and healthy controls for each of the ROIs studied. To determine imaging biomarkers for early PD diagnosis, we generated receiver-operating characteristics (ROC) analysis to identify and predict early PD. P-values less than 0.05 was considered statistically significant.

## 3. Results

### 3.1 Demographic Characteristics of Participants

Demographic and clinical data are depicted in Table 1.

**Table 1.**
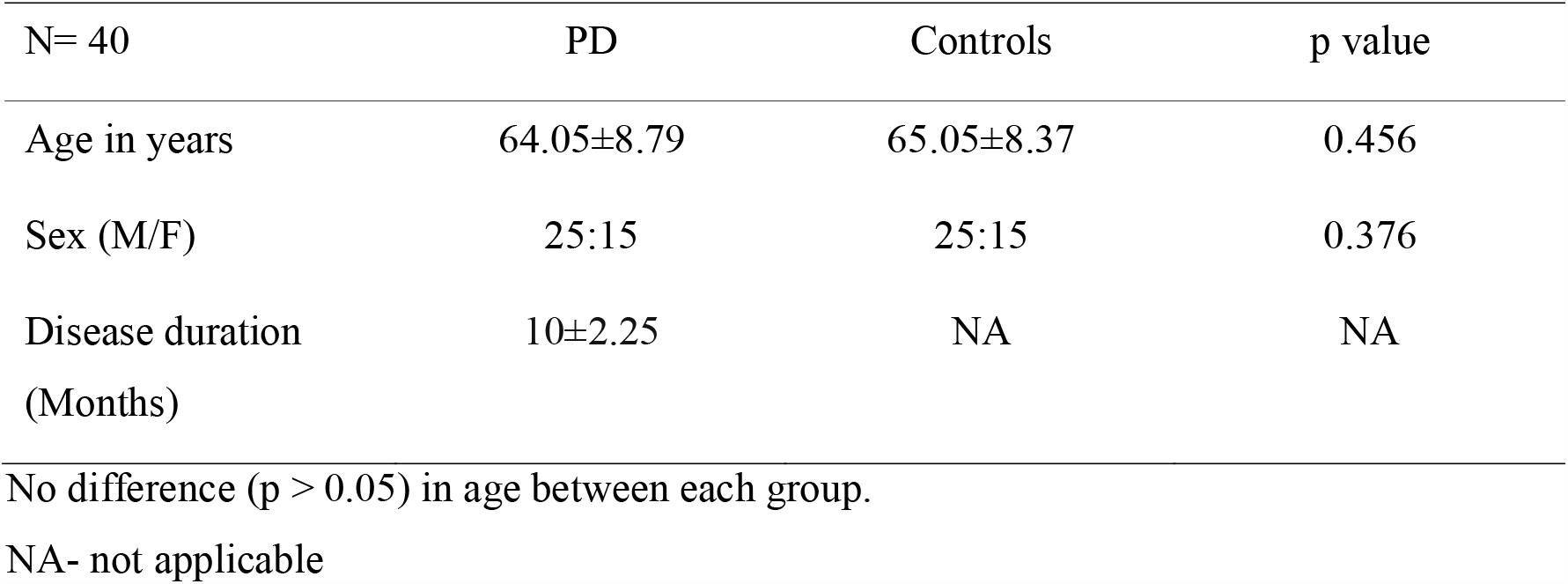
Demographic and clinical data of subjects.

| N= 40 | PD | Controls | p value |
| --- | --- | --- | --- |
| Age in years | 64.05±8.79 | 65.05±8.37 | 0.456 |
| Sex (M/F) | 25:15 | 25:15 | 0.376 |
| Disease duration<br>(Months) | 10±2.25 | NA | NA |

No difference ( $p > 0.05$ ) in age between each group.
NA- not applicable

### 3.2 Diagnostic performance of diffusion tensor imaging

Mean FA values of the brain white and grey matter regions are summarized in Table 2. The highest FA values were in the splenium of the corpus callosum close to 0.70 and the lowest FA values about 0.40 in the caudate nucleus in the PD group compared to control group which showed the highest and lowest FA values of 0.69 at the splenium of the corpus callosum and 0.45 at the caudate nucleus respectively.

**Table 2.** Mean and standard deviation of FA values in PD and control group.

| Sr.<br>No | Regions of the brain | FA @ Controls<br>(Mean ± SD) | FA @ PD<br>(Mean ± SD) |
| --- | --- | --- | --- |
| 1 | Genu of corpus callosum | 0.559±0.86 | 0.660 ±0.07 |
| 2 | Body of corpus callosum | 0.566±0.11 | 0.626±0.11 |
| 3 | Splenium of corpus callosum | 0.693±0.13 | 0.700±0.06 |
| 4 | Centrum Semiovale | 0.529±0.06 | 0.557±0.06 |
| 5 | Pons | 0.586±0.04 | 0.608±0.05 |
| 6 | Caudate Nucleus | 0.454±0.14 | 0.409±0.09 |
| 7 | Putamen | 0.530±0.07 | 0.469±0.08 |
| 8 | Thalamus | 0.508±0.07 | 0.514± 0.08 |
| 9 | Substantia nigra | 0.607±0.07 | 0.422±0.01 |
| 10 | Cerebral peduncles | 0.614±0.05 | 0.645±0.11 |
| 11 | Cerebellar peduncles | 0.508±0.11 | 0.567±0.09 |

### 3.3 Independent sample t-test between PD and control group

We found statistically significant difference between the PD and control group (*P* < 0.05) in the following regions of the brain: Genu and body of corpus callosum, pons, putamen, substantia nigra and cerebral peduncles as depicted in Table. 3.

**Table 3.** Independent sample t test between PD and control group at the white and grey matter regions of the brain.

| Sr. No | Regions of the brain | p value (FA value comparison between PD and Control group) |
| --- | --- | --- |
| 1 | Genu of corpus callosum | 0.000* |
| 2 | Body of corpus callosum | 0.016* |
| 3 | Splenium of corpus callosum | 0.755 |
| 4 | Centrum Semiovale | 0.082 |
| 5 | Pons | 0.037* |
| 6 | Caudate Nucleus | 0.165 |
| 7 | Putamen | 0.000* |
| 8 | Thalamus | 0.214 |
| 9 | Substantia nigra | 0.000* |
| 10 | Cerebral peduncles | 0.003* |
| 11 | Cerebellar peduncles | 0.409 |

\*- signifies statistically significant data

### 3.4 Receiver operator characteristics curves

ROC analysis was done for the regions of the brain which showed statistically significant difference namely the genu and body of the corpus callosum, pons, substantia nigra, putamen and cerebral peduncles to find out positive predictors in PD group compared to controls as shown in Table. 4. The area under the ROC curve (AUC) is a measure of how well a parameter can distinguish between two diagnostic groups (diseased/controls). A rough guide for classifying the accuracy of a diagnostic test is the traditional academic point system: .90-1 = excellent (A) .80-.90 = good (B) .70-.80 = fair (C) .60-.70 = poor (D).

**Table 4.** ROC curve analysis.

| Predictors | Area under the curve | Cut-off | Sensitivity | Specificity |
| --- | --- | --- | --- | --- |
| Genu of corpus callosum | 0.836 | 0.6075 | 0.8 | 0.85 |
| Body of corpus callosum | 0.688 | 0.5545 | 0.8 | 0.625 |
| Pons | 0.630 | 0.586 | 0.575 | 0.475 |
| Putamen | 0.741 | 0.498 | 0.750 | 0.725 |
| Substantia Nigra | 1.0 | 0.484 | 1.000 | 1.000 |
| Cerebral Peduncles | 0.741 | 0.719 | 0.75 | 0.70 |

## 4. Discussion

### 4.1 Substantia nigra and FA values

ROI analysis showed FA values in the substantia nigra were statistically significant and lower in the PD group compared to the control group. Reduced FA values in the substantia nigra of PD patients are a direct effect of loss of neurons. This justifies that the PD group had grey matter alterations which is in line with a systematic review conducted in 2013 and other studies conducted worldwide [19-21]. Our results showed that the FA values in the substantia were reduced in PD patients, suggesting that damage to this structure is directly related to changes in PD consistent with previous studies [22-27]. It has been postulated that damage to the brain results in axonal loss which in turn is associated with changes in FA values.

### 4.2 Putamen and FA values

Lower FA values at the putamen obtained in the study may be used in early prediction of PD. A similar study also quoted the importance of putaminal diffusivity with disease progression in PD with increase in FA values and involvement of the brain grey matter [28]. So, in early PD putamen FA values tend to be on the lower side as stated in the current study findings but as the disease progresses, FA values may tend to increase. Iron deposition in the substantia nigra is observed in PD cases, where iron levels increases FA values at the putamen. This should also be taken into consideration while interpreting FA at the putamen in the clinical set up [29-31].

### 4.3 Corpus callosum and FA values

We found statistically significant difference with higher FA in the PD group compared to the control group in the genu and body of the corpus callosum. Generally notable volume loss occurs in the corpus callosum in late stages of PD. Findings of higher FA at the corpus callosum in PD group compared to controls might be a classical feature seen only in early PD cases. Hence, the findings of increased FA in the genu and body of the corpus callosum sounds noteworthy and can be utilized for diagnosis in early PD cases which is a noble finding and not yet reported in literature. As the disease progresses in PD, dementia, lewy body appearance and Alzheimer’s disease come into play which in turn may cause FA value to reduce as the total volume of the corpus callosum will decrease [32,33]. This trend in corpus callosum has to be sensitively monitored while differentiating early and late PD cases. In general anisotropy depends on the structural arrangement of neurons, compactness and inter-neuronal space. A new advent of a disease, causing disturbance to its normal structure might cause a momentary rise in FA at the corpus callosum but as the disease progresses FA values will tend to fall and be on the lower side.

### 4.4 Cerebral peduncles and FA values

Limited and sporadic studies have been conducted to find the association of cerebral peduncles and FA values and none of them have found any significant results to use cerebral peduncle as a biomarker to predict early PD [34,35]. Some studies have used FA at the cerebral peduncle as a baseline for comparison with disease group. Hence, further studies with larger sample size has to be conducted to evaluate the effectiveness of using cerebral peduncle as a biomarker for detecting early PD.

### 4.5 Pons and FA values

FA in the pons showed statistically significant difference with higher FA in the PD group compared to control group using independent sample ‘t’ test. But roc curve analysis low sensitivity and specificity and hence, did not reveal any positive correlation between PD and control group. Hence, the usefulness of pons as an imaging biomarker remains unclear by the present study findings.

## 5. Conclusion

In early stage parkinsonism, genu and body of the corpus callosum, and cerebral peduncles showed higher FA while lower FA values were seen in the putamen and substantia nigra as compared to healthy controls. Using a ROI approach, FA measures of the substantia nigra seems to be the most appropriate biomarker for differentiating early PD from healthy controls. We consider FA at the putamen as an early predictor of PD. Corpus callosum can be utilized for diagnosis in early PD cases which is a noble finding and not yet reported in literature.

## Data Availability

Availability of data and material- The data collected in the current study comprises of screenshot images taken after obtaining FA values of PD patients and healthy controls using diffusion tensor imaging fibre track software. The data will be shared on request as per the Kasturba Medical hospital ethical committee patient privacy guidelines. The datasets used and/or analysed during the current study is available from the corresponding author on reasonable request.

## Compliance with ethical standard

### Ethical approval

The study protocol followed was reviewed and approved by the Research Committee of Manipal College of Health Profession and Manipal Academy of Higher Education, and ethical clearance was also obtained by Kasturba Medical College and Hospital, MAHE, Manipal

### Informed consent

A detailed explanation about the study was given by the principal investigator after which they provided consent for publication. All the patients included in this research gave written informed consent to publish the data contained within this study.

## ‘Declarations’

### Ethics approval and consent to participate

The study protocol followed was reviewed and approved by the Research Committee of Manipal College of Health Profession and Manipal Academy of Higher Education, and ethical clearance was also obtained by Kasturba Medical College and Hospital, MAHE, Manipal. A detailed explanation about the study was given by the principal investigator after which they provided consent for publication. All the patients included in this research gave written informed consent to publish the data contained within this study.

### Compliance with ethical standards

All ethical standards were maintained.

### Ethics reference number

IEC 407/2014

### Availability of data and material

The data collected in the current study comprises of screenshot images taken after obtaining FA values of PD patients and healthy controls using diffusion tensor imaging fibre track software. The data will be shared on request as per the Kasturba Medical hospital ethical committee patient privacy guidelines. The datasets used and/or analysed during the current study is available from the corresponding author on reasonable request.

### Authors’ contributions

RK conceptualized the study. RK, PK and SN have given inputs in study design. RK collected the data. RK analysed the data and wrote the first draft of manuscript and all co-authors contributed in critical review of data analysis and manuscript writing. RK will act as guarantor for this paper. All authors have read and approved the manuscript.

## Acknowledgements

Not applicable

